# Opaganib in COVID-19 pneumonia: Results of a randomized, placebo-controlled Phase 2a trial

**DOI:** 10.1101/2021.08.23.21262464

**Authors:** Kevin L. Winthrop, Alan W. Skolnick, Adnan M. Rafiq, Scott H. Beegle, Julian Suszanski, Guenther Koehne, Ofra Barnett-Griness, Aida Bibliowicz, Reza Fathi, Patricia Anderson, Gilead Raday, Gina Eagle, Vered Katz Ben-Yair, Harold S. Minkowitz, Mark L. Levitt, Michael S. Gordon

## Abstract

**Background:** Opaganib, an oral sphingosine kinase-2 inhibitor with antiviral and anti-inflammatory properties, was shown to inhibit SARS-CoV-2 replication *in vitro*. We thus considered that opaganib could be beneficial for moderate to severe COVID-19 pneumonia. The objective of the study was to evaluate the effect of opaganib on supplemental oxygen requirements, time to hospital discharge and its safety in COVID-19 pneumonia hospitalized patients requiring supplemental oxygen.

**Methods:** This Phase 2a, randomized, double-blind, placebo-controlled study was conducted between July and December 2020 in eight sites in the USA. Forty-two enrolled patients received opaganib (n=23) or placebo (n=19) added to standard of care for up to 14 days and were followed up for 28 days after their last dose of opaganib/placebo.

**Results:** The relative decrease in total supplemental oxygen requirement from baseline to Day 14 was 61.6% in the opaganib versus 46.7% in the placebo arms. By Day 14, 50.0% of patients in the opaganib and 22.2% in the placebo group no longer required supplemental oxygen for at least 24 hours, while 86.4% and 55.6%, respectively, were discharged from hospital. The incidence of ≥ Grade 3 treatment-emergent adverse events was 17.4% and 33.3% in the opaganib and placebo groups, respectively. Three deaths occurred in each group.

**Conclusions:** In this proof-of-concept study, hypoxic, hospitalized patients receiving oral opaganib required less supplementary oxygen and had earlier hospital discharge, with no safety concerns arising. These findings support further evaluation of opaganib in this population.

**Summary:** Upon receiving opaganib, patients with COVID-19 pneumonia who were hospitalized and required supplemental oxygen showed symptomatic clinical improvement compared to placebo, with less supplemental oxygen requirement, resulting in earlier hospital discharge, and no safety concerns arising.

## INTRODUCTION

Acute pneumonia due to Severe Acute Respiratory Syndrome Coronavirus-2 (SARS-CoV-2) infection leading to coronavirus disease 2019 (COVID-19) can be life-threatening. Typical symptoms of SARS-CoV-2 infection include fever, fatigue, dry cough, shortness of breath, but can evolve to acute respiratory distress syndrome. Among patients with COVID-19, 80% present with symptoms of mild to moderate severity, with or without pneumonia, while 5-14% develop serious or critical respiratory disease, requiring intensive care unit support (1–3). The mortality rate of COVID-19 ranges from 0.1 to 19.7%, depending on country of origin, socioeconomic background, comorbidities such as diabetes and cardiovascular disease, and disease severity (4–8). These comorbidities and advanced age (especially > 65 years) increase the risk of mortality (9, 10).

Hospitalized COVID-19 patients often require supplemental oxygen to maintain adequate oxygenation (3, 9). Some pharmacological agents (corticosteroids and remdesivir) have been widely used as standard of care (SoC), with remdesivir first approved in the USA for emergency use in hospitalized COVID-19 patients in May 2020 (11).

Opaganib (RedHill Biopharma) is a first-in-class orally available, small molecule, selective sphingosine kinase-2 (SK-2) inhibitor with antiviral and anti-inflammatory properties (12). Multiple ongoing clinical trials are evaluating opaganib as therapy for cancer. The antiviral effect of opaganib was demonstrated in various preclinical studies. Reid and colleagues showed that decreased expression or pharmacologic inhibition of SK-2 significantly inhibited Chikungunya virus replication (13). In a work by Xia *et al*., opaganib was shown to reduce fatality rates from influenza A infection in mice (14). Moreover, in a preliminary study using an *in vitro* model of human bronchial tissue, we recently demonstrated that opaganib has potent antiviral activity against SARS-CoV-2 (15). We hypothesized that the antiviral and anti-inflammatory effects of opaganib would be beneficial for the treatment of SARS-CoV-2 infection, particularly for patients with moderate to severe COVID-19 pneumonia requiring supplemental oxygen. In a small cohort of patients with severe COVID-19 pneumonia, where opaganib was offered through compassionate use, it appeared to improve clinical and laboratory parameters, and was safe and well tolerated (16).

This Phase 2a proof-of-concept study was designed to evaluate the effect of opaganib when added to SoC in hospitalized patients with COVID-19 pneumonia who required supplemental oxygen. Efficacy endpoints evaluated the change in oxygen requirements, clinical improvement parameters, and safety assessments.

## METHODS

### Design

This was a Phase 2a, proof-of-concept, multi-center, randomized, double-blind, parallel arm, placebo-controlled study conducted in eight sites in the USA. Eligible patients received opaganib or matching placebo twice daily added to SoC (per regional, institutional/physician practices). Eligible participants were adults (18-80 years inclusively), with SARS-CoV-2 infection determined by polymerase-chain-reaction (PCR) analysis in a nasopharyngeal sample, and pneumonia determined by chest X-ray. The full list of eligibility criteria is provided in the supplementary data (Section-1).

### Intervention

Participants were randomized 1:1 to receive 2 × 250 mg opaganib oral capsules or matching placebo every 12 hours for up to 14 days (Days 1-14). This dose had been determined in a Phase 1 clinical study in oncology patients as the maximum safe and tolerable dose (17). Patients discharged prior to/at Day 10 completed only 10 days of treatment. After their last study drug dose, participants were followed for up to 28 days (safety follow-up). Treatment assignments remained blinded to the patients, investigators, hospital staff, and sponsor (RedHill Biopharma). The clinical trial (NCT 04414618) was performed in accordance with the Declaration of Helsinki (1964), as revised most recently in Seoul (2008), US Food and Drug Administration (FDA) regulations and the ICH Guideline for Good Clinical Practice, E6(R1) and local rules and regulations.

### Outcomes

Given the exploratory nature of this proof-of-concept study, we sought to evaluate several clinically meaningful outcomes. We designed a primary outcome measure comparing total supplemental oxygen requirements from baseline to day 14 (area-under-the-curve [AUC]) using daily oxygen flow measurements. Primary endpoint details are provided in the supplementary data (Section-2), together with a complete list of secondary, post-hoc, safety and exploratory endpoints. Presented herein are the following clinically meaningful secondary outcomes: (a) time to 50% reduction from baseline in supplemental oxygen requirement by Day 14, (b) percentage of patients no longer receiving supplemental oxygen for at least 24 hours by Day 14, (c) time to intubation and mechanical ventilation by Day 14, (d) mortality by Day 30; post-hoc outcomes: (a) percentage of patients no longer receiving supplemental oxygen for at least 24 hours by Day 14 per SoC regimen, (b) time to intubation and mechanical ventilation by safety follow-up, (c) mortality by safety follow-up, (d) incidence of hospital discharge by Day 14 and by safety follow-up, (e) time to ≥2 points improvement in the WHO Ordinal Scale (18) by Day 14; safety outcome: incidence of treatment-emergent adverse events (TEAEs) and serious TEAEs over the safety follow-up period.

### Statistics

As the sample size (∼40 patients) was not chosen for statistical consideration, efficacy interpretation is based on numerical comparisons of descriptive statistics. Statistical methodology details are provided in the supplementary data (Section-3). Baseline characteristics are presented for the Intent-to-Treat (ITT) population (all randomized patients). Efficacy endpoints were analyzed for the modified ITT (mITT) population (patients who took at least one dose of study drug). Safety evaluations were performed for the safety population (patients who took at least one dose of study drug).

## RESULTS

A diagram of the study flow is presented in Figure 1. The ITT population comprised of 42 eligible patients, enrolled July-November 2020. Of these, 23 patients were randomized to receive opaganib and 19 patients were randomized to receive placebo (randomization details are provided in the supplementary data, Section-4). One patient in each group did not receive study drug and was excluded from the safety and mITT populations. One patient randomized to placebo was mistakenly given opaganib on Days 10 and 11.

**Figure 1.**
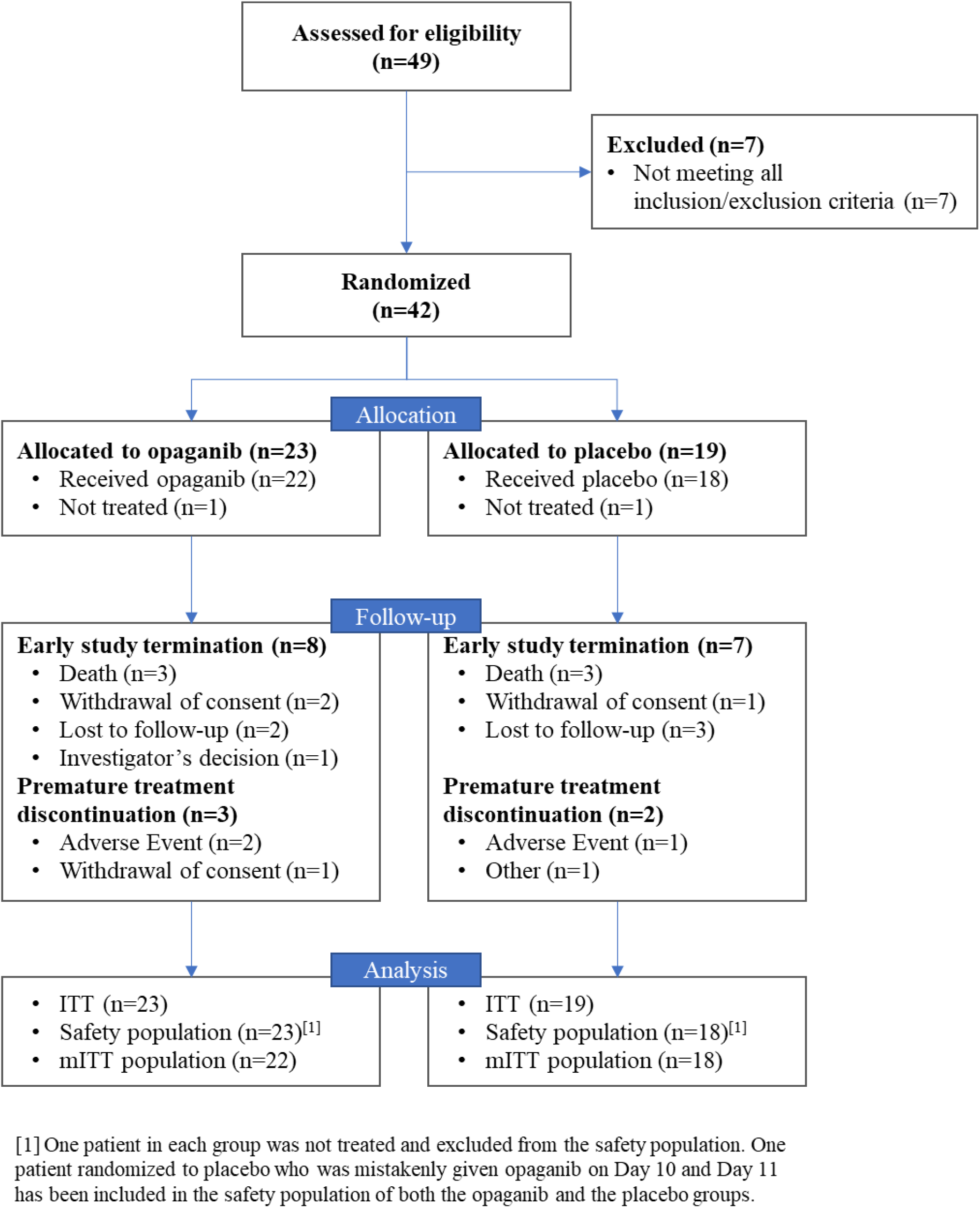
Study flow diagram. Abbreviations: ITT – Intent-to-Treat population; mITT – modified Intent-to-Treat population.

### Demographic and other baseline characteristics

The main demographic and baseline characteristics of patients in the ITT population are presented in Table 1. Most patients were male (64.3%) with an overall median age of 58.0 years. Randomization resulted in a difference between the opaganib and placebo arms in median age (years; 52.0 and 61.0, respectively), and median baseline oxygen requirement (L/min; 6.0 and 10.5, respectively). The median time (days) from diagnosis to hospitalization was 0.0 and the median time from diagnosis to randomization was 4.0, and similar for both groups.

**Table 1:** Main patient demographic and baseline characteristics in the opaganib and placebo groups and overall.

| Parameter (ITT population) | Opaganib<br>(N=23) | Placebo<br>(N=19) | Overall<br>(N=42) |
| --- | --- | --- | --- |
| Age (years), median (range) | 52.0 (29, 80) | 61.0 (35, 80) | 58.0 (29,80) |
| Age (years), n (%) |  |  |  |
| <70 | 20 (87.0) | 15 (78.9) | 35 (83.3) |
| ≥70 | 3 (13.0) | 4 (21.1) | 7 (16.7) |
| Gender, n (%) |  |  |  |
| Male | 16 (69.6) | 11 (57.9) | 27 (64.3) |
| Female | 7 (30.4) | 8 (42.1) | 15 (35.7) |
| Ethnicity, n (%) |  |  |  |
| Hispanic or Latino | 12 (52.2) | 8 (42.1) | 20 (47.6) |
| Race, n (%) |  |  |  |
| White | 18 (78.3) | 15 (78.9) | 33 (78.6) |
| American Indian or Alaska Native | 1 (4.3) | 1 (5.3) | 2 (4.8) |
| Asian | - | 1 (5.3) | 1 (2.4) |
| Black or African American | 3 (13.0) | 2 (10.5) | 5 (11.9) |
| Other | 1 (4.3) | - | 1 (2.4) |
| Smoking status, n (%) |  |  |  |
| Never | 20 (87.0) | 14 (73.7) | 34 (81.0) |
| Former | 2 (8.7) | 4 (21.1) | 6 (14.3) |
| Current | 1 (4.3) | 1 (5.3) | 2 (4.8) |
| HbA1c (%) at screening, n (%) |  |  |  |
| <6.5 | 13 (56.5) | 10 (52.6) | 23 (54.8) |
| ≥6.5 | 10 (43.5) | 9 (47.4) | 19 (45.2) |
| Weight at baseline (Kg), median (range) <sup>a</sup> | 104.33 (74.8, 163.3) | 79.00 (55.3, 140.0) | 96.76 (55.3, 163.3) |
| Oxygen requirement at baseline (L/min) |  |  |  |
| n | 22 | 18 | 40 |
| Median (min, max) | 6.0 (0, 40) | 10.5 (1, 55) | 6.0 (0, 55) |
| Time from symptoms onset to hospitalization (days), median (range) <sup>b</sup> | 7.0 (-4, 39) | 6.5 (0, 11) | 7.0 (-4, 39) |
| Time from symptoms onset to randomization (days), median (range) <sup>b</sup> | 10.0 (4, 41) | 9.0 (2, 18) | 9.0 (2, 41) |
| Time from diagnosis to hospitalization (days), median (range) | 0.0 (-5, 20) | 0.0 (-1, 14) | 0.0 (-5, 20) |
| Time from diagnosis to randomization (days), median (range) | 4.0 (1, 33) | 4.0 (2, 18) | 4.0 (1, 33) |

Abbreviations: HbA<sub>1c</sub> – hemoglobin A<sub>1c</sub>
<sup>a</sup> Two patients in each group had missing information on weight at baseline, resulting in n=21 patients in the opaganib group and n=17 patients in the placebo group.
<sup>b</sup> One patient in the placebo group had missing information on symptoms onset, resulting in n=18 patients in the placebo group.

Remdesivir and/or high dose corticosteroids (dexamethasone, prednisone, and methylprednisolone) comprised known effective SoC for COVID-19 and were co-administered with opaganib in 22 (95.7%) patients, and with placebo in all 18 (100%) patients. This included remdesivir in 10 (43.5%) patients on opaganib and nine (50.0%) on placebo, and high dose corticosteroids in 21 (91.3%) and all 18 (100%) patients, respectively.

### Treatment compliance

Compliance was assessed in a subset of the safety population comprising of 21 patients on opaganib and 18 patients on placebo (further details are provided in the supplementary data, Section-5). For two patients on opaganib, compliance was not defined: for the patient randomized to placebo who took opaganib on one day, the expected number of opaganib doses was zero; the second patient was lost to follow up, thus compliance could not be assessed. The mean (±standard deviation) compliance was 90.05% (±21.523) in the opaganib and 98.41% (±3.917) in the placebo groups, respectively.

### Total supplemental oxygen requirement using the maximal daily oxygen flow over 14 days

The relative benefit derived for each group was calculated as 61.6% (−770/-1250) for opaganib (n=21) and 46.7% (−583.6/-1250) for placebo (n=18) (Figure 2; details on the relative benefit calculation are provided in the supplementary data, Section-2). This was a post-hoc analysis to the pre-defined primary endpoint; as more information on the course of COVID-19 pneumonia became available, these endpoints, quantifying total supplemental oxygen required over time (across various devices), proved to be of less clinical value than initially assessed.

**Figure 2:**
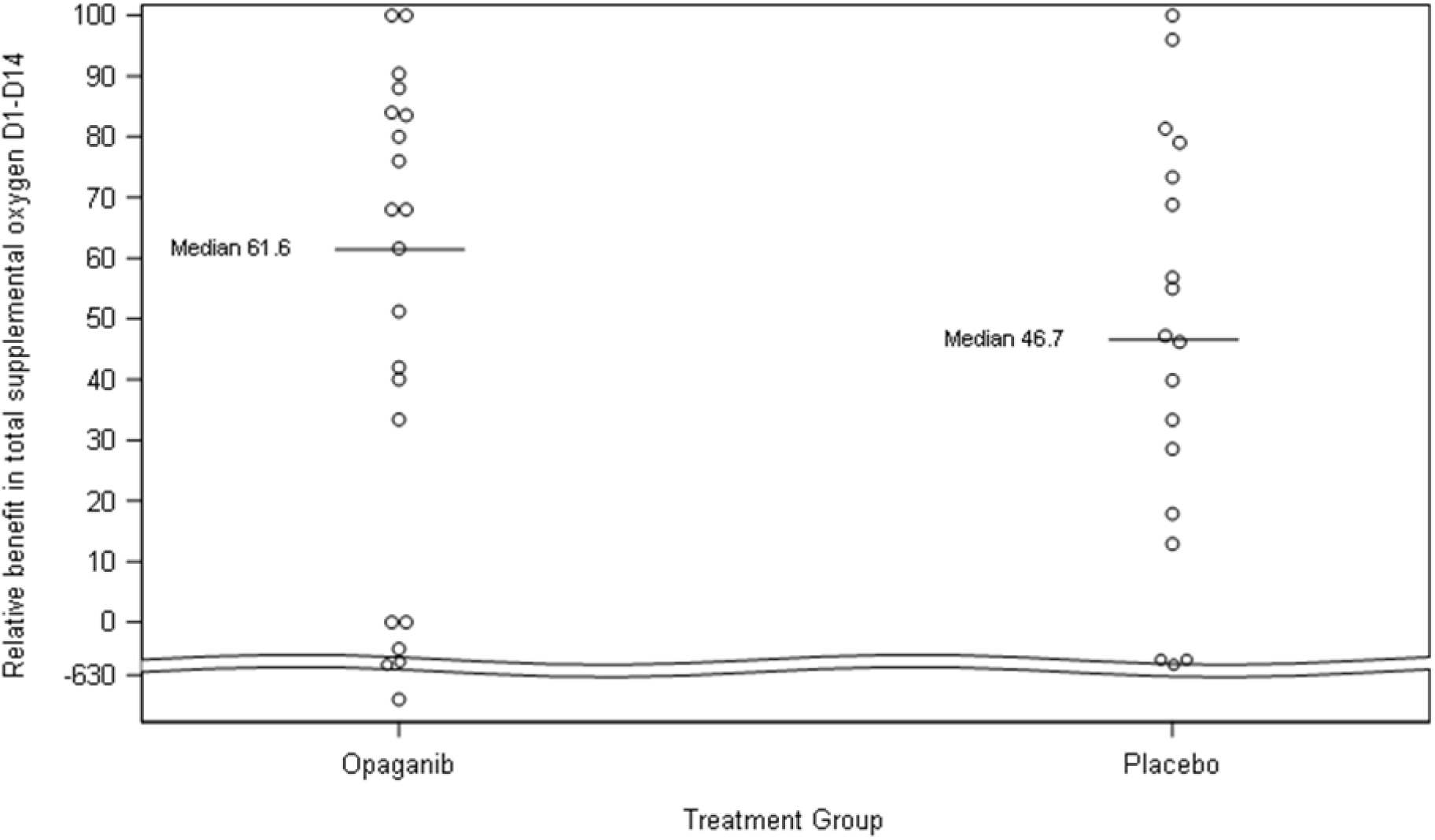
Dot plot of relative improvement in total supplemental oxygen from Day 1 to Day 14 of treatment in the opaganib and placebo groups in the mITT population. The total number of patients in the opaganib group is 21 (rather than 22 in the modified Intent-to-Treat [mITT] population) because one patient did not require oxygen at baseline and was therefore excluded from this analysis.

### Requirement of supplemental oxygen by Day 14

The Kaplan-Meier analysis for no longer requiring supplemental oxygen for at least 24 hours by Day 14 showed a higher estimated cumulative incidence in patients on opaganib (50%) compared to those on placebo (22.2%) (Figure 3, Table 2).

**Figure 3:**
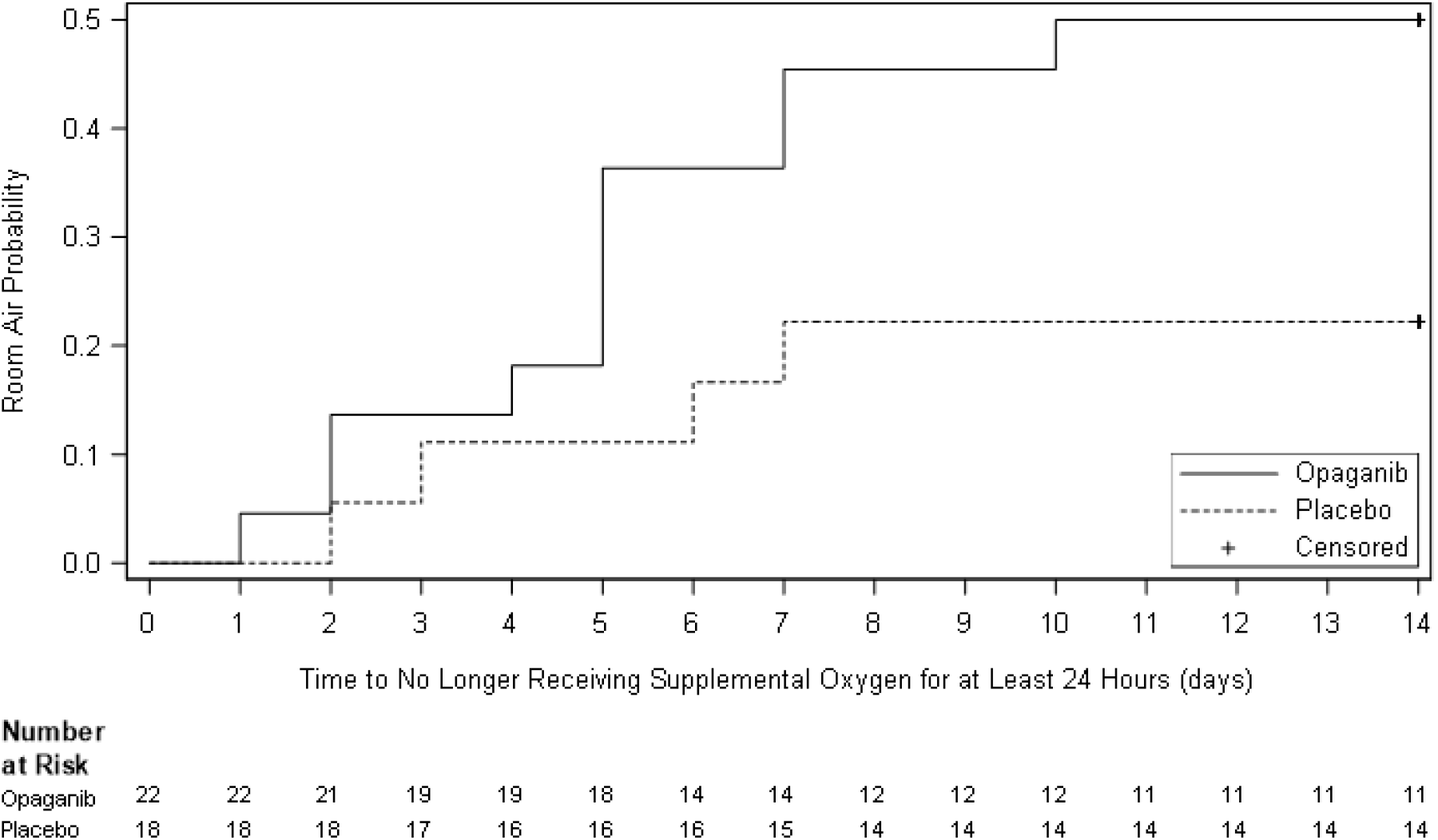
Kaplan-Meier curve of time (days) to no longer receiving supplemental oxygen for at least 24 hours by Day 14 (mITT population). Death was censored at Day 14 as the worst outcome for this endpoint.

**Table 2:** Kaplan-Meier estimate of the cumulative incidence of no longer receiving supplemental oxygen for at least 24 hours by Day 14 per standard of care regimen of interest in the opaganib and placebo groups. Death was censored at 14 Days.

| **Standard of Care Regimen Group (mITT population) <sup>a</sup>** | **Opaganib**<br>**(N=22)** | **Placebo**<br>**(N=18)** |
| --- | --- | --- |
| Overall (N) | 22 | 18 |
| Number of events <sup>b</sup>, n (%) | 11 (50.0) | 4 (22.2) |
| Cumulative incidence at Day 14 <sup>c</sup> | 50.00 | 22.22 |
| Treated with combination of remdesivir and high dose corticosteroids (N) | 9 | 9 |
| Number of events <sup>b</sup>, n (%) | 4 (44.4) | 2 (22.2) |
| Cumulative incidence at Day 14 <sup>c</sup> | 44.44 | 22.22 |
| Treated only with remdesivir (N) | 1 | - |
| Number of events <sup>b</sup>, n (%) | - | - |
| Cumulative incidence at Day 14 <sup>c</sup> | 0.00 | - |
| Treated only with high dose corticosteroids (N) | 11 | 9 |
| Number of events <sup>b</sup>, n (%) | 6 (54.5) | 2 (22.2) |
| Cumulative incidence at Day 14 <sup>c</sup> | 54.55 | 22.22 |
| Neither Standard of Care (N) | 1 | - |
| Number of events <sup>b</sup>, n (%) | 1 (100) | - |
| Cumulative incidence at Day 14 <sup>c</sup> | 100.00 | - |

<sup>a</sup> The number of patients treated with specific standard of care regimen is presented.
<sup>b</sup> Event is defined as no longer receiving supplemental oxygen for at least 24 hours.
<sup>c</sup> The cumulative incidence of patients was estimated with the Kaplan–Meier method.

The subgroup analysis based on SoC regimen showed consistent findings across all SoC sub-groups (Table 2). Specifically, the estimated cumulative incidence of no longer receiving supplemental oxygen by Day 14 in patients receiving opaganib on top of remdesivir in combination with high dose corticosteroids (44.4%), and in those receiving opaganib on top of high dose corticosteroids only (54.6%), was consistent with that observed in the overall opaganib group, and higher than in the respective subgroups receiving placebo on top of the SoC (44.4% versus 22.2%, and 54.6% versus 22.2%, respectively).

The median Kaplan-Meier estimated time to 50% reduction from baseline in supplemental oxygen requirement based on oxygen flow (L/min) Day 1-Day 14 was 5 days in the opaganib versus 8 days in the placebo group. The Kaplan-Meier estimated cumulative incidence of this event by Day 14 was 81% in the opaganib group compared to 66.7% in the placebo group.

### Intubation and mechanical ventilation, and mortality during the course of the study and at the end of follow-up

The Kaplan-Meier estimated cumulative incidence of intubation and mechanical ventilation at Day 14 was 9.6% in the opaganib group and 11.1% in placebo. There were two intubation and mechanical ventilation events in each group, and no subsequent intubations and mechanical ventilations through the end of safety follow-up.

Three patients on opaganib and two patients on placebo died by Day 30. The Kaplan-Meier estimated cumulative incidence of death by Day 30 was 15.0% in the opaganib and 11.9% in the placebo group. An additional death occurred in the placebo group by the end of safety follow-up. The Kaplan-Meier estimated cumulative incidences of death by safety follow-up were 15.0% and 19.2% in the opaganib and placebo groups, respectively.

### Hospital discharge

The Kaplan-Meier estimated cumulative incidence of hospital discharge was 68.2% in the opaganib and 50.0% in the placebo groups by Day 7; and 86.4% and 55.6%, respectively by Day 14 (Figure 4).

**Figure 4:**
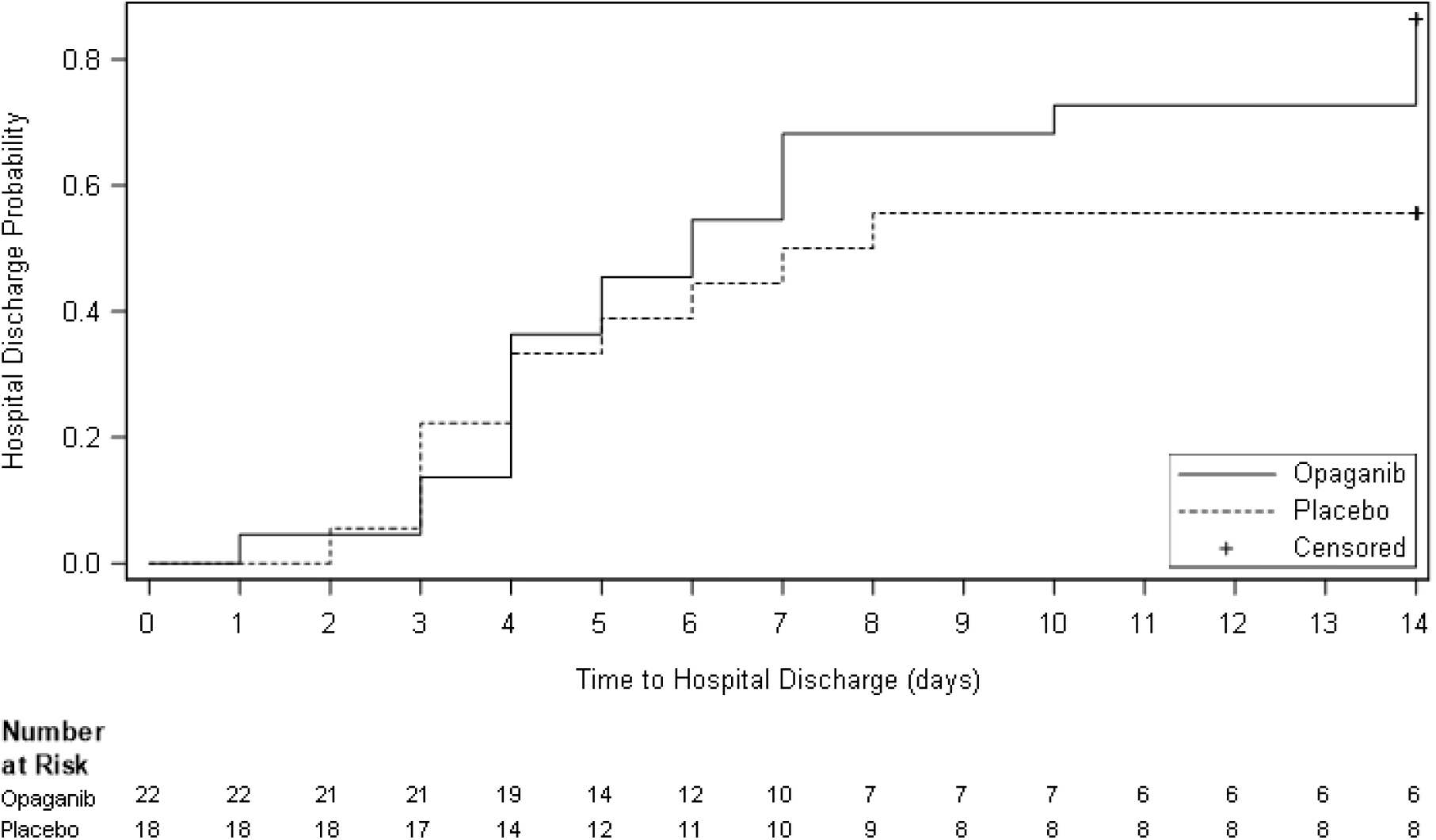
Kaplan-Meier curve of time to hospital discharge by Day 14 in the opaganib and placebo groups (mITT population). Death was censored at Day 14 as the worst outcome for this endpoint.

In a related post-hoc analysis, we derived the time to a 2-point improvement in the WHO Ordinal Scale (18) (details are provided in Table S1). All patients, except for one who did not require supplemental oxygen at baseline, started the treatment while hospitalized and receiving supplemental oxygen (score 4 or 5). Patients discharged from hospital had to reach a score of at most 2, ie, show improvement of at least two points on the WHO ordinal scale. The Kaplan-Meier estimated cumulative incidence of ≥2-point improvement on the WHO Ordinal Scale by Day 14 was 81.8% in the opaganib group, and 55.6% in the placebo group (Table 3). Additionally, the estimated median time (days) to achieving this improvement was shorter in opaganib (6.0 days) compared to placebo (7.5).

**Table 3:** Time to two point improvement on the WHO Ordinal Scale by Day 14 (mITT population).

| **mITT population** | **Opaganib**<br>**(N=22)** | **Placebo**<br>**(N=18)** |
| --- | --- | --- |
| Number with two-point improvement by Day 14 (%) | 18 (81.8) | 10 (55.6) |
| Number of censored by Day 14 (%) | 4 (18.2) | 8 (44.4) |
| Kaplan-Meier cumulative incidence <sup>a</sup> (95% CI) | 81.82 (63.71 - 94.32) | 55.56 (34.88 - 78.42) |
| Kaplan-Meier median estimate (95% CI) (days) | 6.00 (4.00 – 14.00) | 7.50 (4.00 – NA) |

Abbreviations: CI – Confidence interval; NA – Not achieved
<sup>a</sup> The cumulative incidence of patients was estimated with the Kaplan–Meier method.

### Treatment emergent adverse events

The incidence of TEAEs was similar between groups (52.2%, in the opaganib group versus 50.0% in the placebo group; Table 4). Generally, each unique TEAE term was reported singly by one patient (either in the opaganib or placebo group). The only TEAEs reported by two patients (8.7% in the opaganib group), and with a higher incidence in the opaganib group than in placebo, were hypokalemia (reported in one patient on placebo) and diarrhea (not reported in patients on placebo).

**Table 4:**
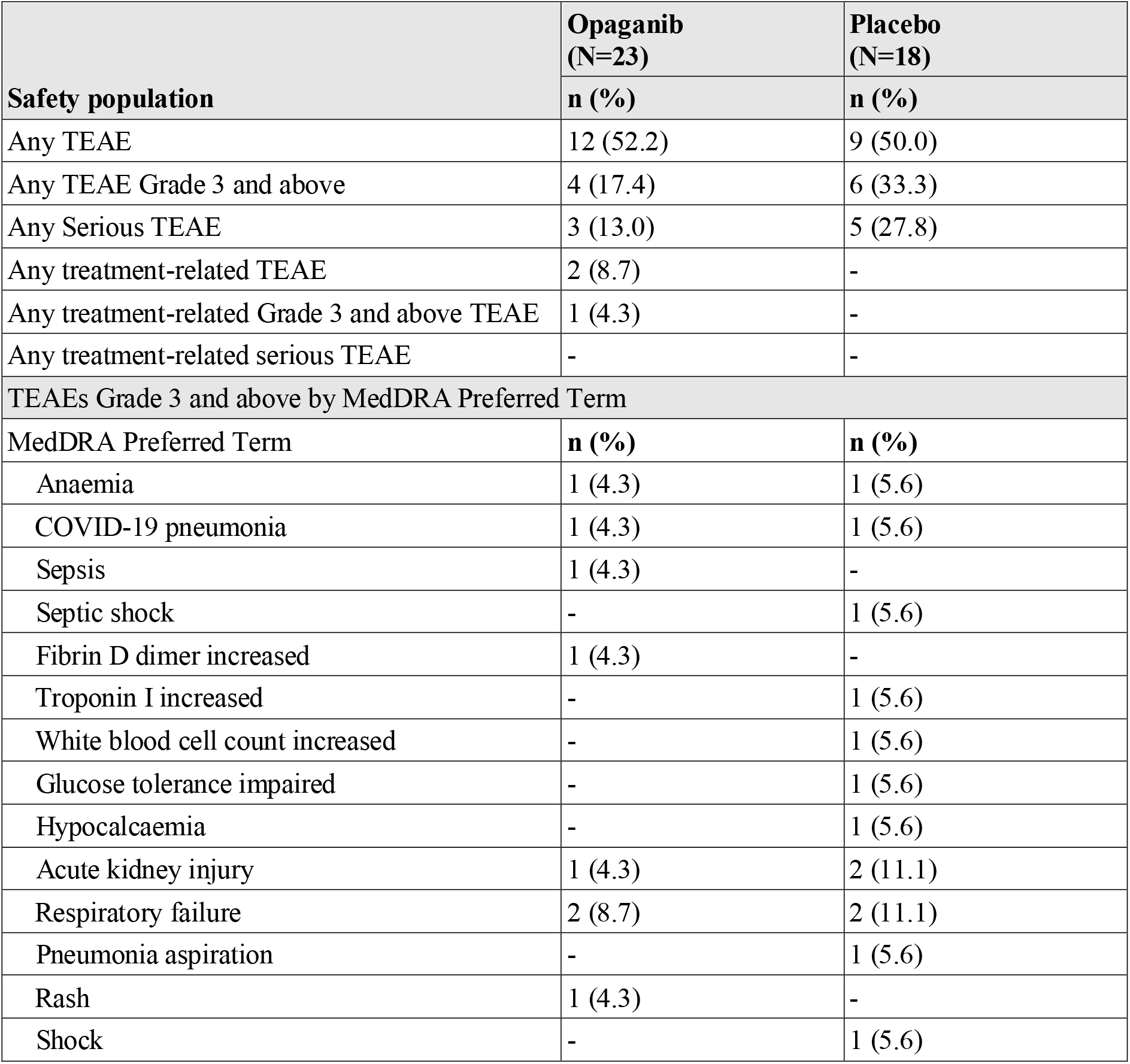
Treatment emergent adverse events (TEAEs) by Medical Dictionary for Regulatory Activities (MedDRA) Preferred Term in the opaganib and placebo groups (safety population)

Serious TEAEs were reported in 13.0% of patients on opaganib and 27.8% patients on placebo. TEAEs of Grade 3 and above were reported in 17.4% of patients on opaganib and 33.3% of patients on placebo, with no TEAEs of Grade 3 and above reported at a higher frequency in the opaganib group compared to placebo (Table 4). There were no reported TEAEs resulting in dose reductions. TEAEs leading to treatment withdrawal were reported in 17.4% of the patients in the opaganib group, and 5.6% of the patients in the placebo group. Three death cases were reported in each group.

## DISCUSSION

This proof-of-concept study explored the effectiveness and safety of oral opaganib in hospitalized patients with COVID-19 pneumonia requiring supplemental oxygen. Results suggest clinical improvement as measured by reduced need for supplemental oxygen, improved WHO level in the scale for clinical improvement and in time to discharge. Opaganib’s safety profile was not materially different than that of placebo.

Several lines of preliminary evidence suggested that opaganib may be particularly relevant to COVID-19 therapy. Prior to the onset of the COVID-19 pandemic, opaganib had demonstrated antiviral and anti-inflammatory effects in several preclinical models, including amelioration of *Pseudomonas aeruginosa*-induced lung injury and reduction of fatality rates from influenza infection (19, 20); inhibition of host inflammatory responses in several disease models (21, 22); and exertion of various effects on the immune system (23). Opaganib has since been shown to have potent antiviral activity against SARS-CoV-2 in a preclinical study using an *in vitro* model of human bronchial tissue (15) and was associated with favorable outcomes when offered through compassionate use to patients with moderate to severe COVID-19 pneumonia requiring supplemental oxygen via high-flow nasal cannulae (16). As opaganib targets a host cell factor rather than the virus directly, development of resistance is less likely. This is of particular interest in SARS-CoV-2 infection, given its rapidly emerging mutations and viral strains.

The antiviral agent remdesivir has been approved for treatment in certain stages of COVID-19; however, it is administered intravenously and lacks anti-inflammatory properties (5), while opaganib is an oral agent with both antiviral and anti-inflammatory properties. Moreover, opaganib is stable at room temperature for several years (RedHill Biopharma Ltd., unpublished data). These factors support opaganib as a much needed, practical and convenient potential treatment of COVID-19 pneumonia.

The current study was designed to primarily assess the change in total supplemental oxygen requirements per patient during the 14-day course of treatment. The pre-specified analysis used absolute change and a post-hoc analysis utilized percent change. This post-hoc analysis demonstrated an apparent difference, with less total supplemental oxygen requirement for patients on opaganib compared to those on placebo. However, as the pandemic progressed, with more information on the course of COVID-19 pneumonia becoming available, it became evident that the clinical interpretation of this endpoint was limited, given high variability in baseline values and in devices used to supply oxygen. Additionally, since most patients were discharged before Day 14, certain pre-defined secondary efficacy analyses based on 14 days of in-patient data collection could not be performed. In fact, as a higher proportion of patients on opaganib were discharged from the hospital before Day 14 compared to those on placebo, missing data is not balanced, and likely to introduce a bias to these secondary efficacy analyses. As improvement in supplemental oxygen requirement had become a commonly used criterion for hospital discharge in COVID-19 patients (24), our pre-defined secondary analysis of no longer requiring supplemental oxygen for at least 24 hours was the most clinically relevant endpoint. Further post-hoc analyses demonstrated that compared to placebo, 1) the benefit of no longer requiring supplemental oxygen at Day 14 when treated with opaganib was maintained irrespective of the background SoC; 2) hospital discharge was more rapid for patients on opaganib; 3) a derived 2-point improvement in the WHO Ordinal Scale occurred sooner for patients on opaganib. These post-hoc analyses indicated clinical improvement in disease severity, supporting the pre-specified efficacy endpoints. All evaluated secondary and post-hoc efficacy endpoints demonstrated a numerically superior benefit for patients on opaganib compared with those on placebo.

Of note, there was an imbalance in median age between the two arms of the study, with a younger population in the opaganib group (52.0 years) compared to placebo (61.0 years). Additionally, the mean baseline oxygen requirement (L/min) was lower in the opaganib group (6.0) than in placebo (10.5). As younger individuals are expected to have generally better disease outcomes than their older counterparts, we cannot rule out that these imbalances may have impacted the results of the study, contributing to opaganib’s observed benefit in the studied population. However, such potential impact remains hypothetical, as most studies looking at the effect of age on disease severity, report the age of > 65 years as a risk factor.

Opaganib was shown to be well-tolerated in this study, with no new safety signals emerging. The only TEAE of Grade 3 and above reported at a higher frequency in the opaganib group than placebo was a Grade 3 rash in a single patient, that resolved upon stopping study drug. Rash is a known uncommon toxicity of opaganib. Three patients on opaganib and five patients on placebo experienced TESAEs. TEAEs leading to treatment withdrawal were reported in 17.4% of patients on opaganib, and 5.6% of patients on placebo. By the safety follow-up timepoint, there were three deaths recorded in each group, none deemed treatment related. The overall incidence of TEAEs and TESAEs were similar in both treatment groups.

### Limitations

The main limitation of this study is its small sample size, allowing for descriptive analyses rather than meaningful statistical interpretation. A further limitation is the imbalance in age and supplemental oxygen requirement at baseline, a result of the small sample size despite randomization. The clinical interpretation of the primary endpoint was limited, as described in the Discussion, leading to the addition of post-hoc analyses to help with the overall interpretation of the data. An additional limitation of the study is that certain pre-defined secondary endpoints that relied on inpatient data collection could not be assessed due to changing standard of practice leading to earlier discharge of patients than anticipated at the time of protocol development. Furthermore, discharge criteria were somewhat subjective, depending on hospital capacity and regulations. Notably, a greater proportion of patients on opaganib versus placebo were discharged prior to Day 14.

### Conclusions

Taken together, the results of this Phase 2a study suggest a potential benefit of opaganib across various clinically meaningful outcome measures in patients with COVID-19 pneumonia, with no new safety concerns. Based on these findings, further evaluation of opaganib is currently ongoing in larger randomized studies as a potential treatment for moderate-severe COVID-19 pneumonia in hospitalized patients requiring supplemental oxygen (a global Phase 2/3 [NCT04467840] already completed enrollment (15)).

## Data Availability

In general, RedHill Biopharma Ltd. (RedHill) adopts ICMJE requirements regarding data sharing as detailed in its Data Sharing Plan.
In accordance, the following statements apply to the publication titled Opaganib in COVID-19 pneumonia: Results of a randomized, placebo-controlled Phase 2a trial:
Individual deidentified participant data (including data dictionaries) will currently not be shared
Additional, related documents will be not available (e.g., study protocol, statistical analysis plan, etc.)
Will the data become available and for how long? These are currently not available therefore this is not applicable.
Since RedHill currently does not share individual deidentified participant data there are no access criteria wherein data will be shared (including with whom, for what types of analyses, and by what mechanism).

## ACKNOWLEDGMENTS ^a,b^

The authors would like to acknowledge the patients who participated in the study, as well as the dedicated healthcare professionals who cared for them at risk of their own lives. The authors would like to thank Dr Dana Savulescu (Bioforum Group, Ltd.) for her assistance in manuscript editing.

