## Supplementary Data for "Opaganib in COVID-19 pneumonia: Results of a randomized, placebo-controlled Phase 2a trial"

### 1. Participant eligibility

For inclusion into the study, patients were to fulfill all of the following criteria:

- Adult male or female ≥18 to ≤80 years of age
- Proven severe acute respiratory syndrome coronavirus 2 (SARS-CoV-2) infection per real time by polymerase chain reaction (PCR) assay of a pharyngeal sample (nasopharyngeal or oropharyngeal) AND pneumonia defined as radiographic opacities on chest X-ray
- The patient required supplemental oxygen at baseline
- The patient, guardian or legal representative has signed a written Institutional Review Board-approved informed consent
- Male participants with female partners of child-bearing potential agreed to one of the following methods of contraception during the treatment period and for at least one month after the last dose of study drug:
  - Abstinence from penile-vaginal intercourse and agree to remain abstinent
  - Male condom, with female partner using a highly effective contraceptive method
- In addition, male participants had to refrain from donating sperm for the duration of the study and for one month after last dose of study drug.
- Male participants with a pregnant or breastfeeding partner had to agree to remain abstinent from penile-vaginal intercourse or use a male condom during each episode of penile penetration for at least one month after the last dose of study drug.
- Female participants: a female participant was eligible to participate if she was:
  - not pregnant
  - not breastfeeding

- not a woman of child-bearing potential (WOCBP)
- a WOCBP who agreed to use a highly effective method of contraception consistently and correctly during the treatment period and for at least one month after the last dose of study drug

Patients were excluded from the study if during screening they met any of the following criteria:

- Any co-morbidity that could add risk to the treatment in the judgment of the Investigator
- Requiring intubation and mechanical ventilation
- Patient having a “do not intubate” or “do not resuscitate” order
- Oxygen saturation >95% on room air
- Any preexisting respiratory condition that required intermittent or continuous ambulatory oxygen prior to hospitalization
- Patient was, in the Investigator’s clinical judgment, unlikely to survive >72 hours
- Pregnant (positive serum test within three days prior to randomization) or nursing women
- Unwillingness or inability to comply with procedures required in this protocol
- Corrected QT (QTc) interval on electrocardiogram (ECG) >470 ms for females or >450 ms for males, calculated using Friedericia’s formula (QTcF)
- Aspartate aminotransferase (serum glutamic oxaloacetic transaminase) (AST [SGOT]) or Alanine aminotransferase (serum glutamic pyruvate transaminase) (ALT [SGPT]) >5.0 x upper limit of normal (ULN)
- Bilirubin >2.0 x ULN (except where bilirubin increase is due to Gilbert’s Syndrome)
- Serum creatinine >2.0 x ULN
- Absolute neutrophil count <1000 cells/$mm^3$

- Platelet count <75,000/mm$^3$
- Hemoglobin (referred to as HbA1c in the Results) <8.0 g/dL
- Taking medications that are sensitive CYP3A4, CYP1A2, CYP2C9, CYP2C19 or CYP2D6 substrates and have a narrow therapeutic index at the time of screening. These had to be decided in discussion with the Medical Monitor on a case-by-case basis.
- Taking medications that are strong inducers or inhibitors of CYP2D6 and CYP3A4 at the time of screening. These had to be decided in discussion with the Medical Monitor on a case-by-case basis.
- Taking warfarin, apixaban, argatroban or rivaroxaban at the time of screening
- Drug or alcohol abuse at the time of screening
- Participating in a clinical study assessing pharmacological treatments, including anti-viral studies at the time of screening

**2. Study endpoints**

The pre-defined primary efficacy endpoint of the study was to evaluate the effect of opaganib on total supplemental oxygen requirement (area-under-the-curve [AUC] using daily oxygen flow (L/min) measurements for 14 days. For each patient, the AUC was to be calculated using the trapezoidal rule through day 14, subtracting the baseline oxygen requirement at each day. Upon review of the raw data after unblinding, an apparent imbalance among treatment groups in baseline oxygen flow requirement was found, with a median of 6 L/min in the opaganib group versus 10.5 L/min in placebo. This was accompanied by a large variability in these baseline values. Therefore, a post-hoc primary endpoint was added to enhance our ability to assess activity, for which the AUC calculation was based on the daily percent change (reduction or increase) from baseline curve, rather than the absolute change from baseline curve. In this approach, we were able to present relative benefit in total oxygen requirement from baseline through Day 14 with the patients' AUC results calculated relative to the best possible outcome a patient could achieve. The maximal possible AUC benefit of -1250% is achieved if supplemental oxygen is no longer required (100% reduction) as early as Day 2 (the day after baseline) and maintained through Day 14 (derivation was performed as follows, applying the trapezoidal rule: the area of the Day 1 to Day 2 triangle is (0+-100)/2=-50, where subsequent 12 daily rectangles from Day 2 to Day 14 have an area of (-100+-100)/2=-100, each; the sum of these areas is -1250 as the best possible outcome). The relative benefit, calculated for each patient relative to the maximal score of -1250%, could potentially range between any negative benefit (corresponding to increase in supplemental oxygen requirement) to a score of 100%, which corresponds to an optimal result. The median percent change from baseline in the supplemental oxygen requirement AUC over 14 days was -770% for the opaganib group (a median relative benefit -770/-1250=61.6%) versus -583.6% for placebo (a median relative benefit -583.6/-1250=46.7%), showing a numerically greater reduction in total oxygen requirement for opaganib compared to placebo.

The pre-defined secondary efficacy endpoints included the following: (1) the time to 50% reduction from baseline in supplemental oxygen based on oxygen flow in L/min; (2) the proportion of subjects no longer requiring supplemental oxygen for at least 24 hours by Day 14; (3) the proportion of afebrile subjects at Day 14; (4) the time to negative swabs for SARS-CoV-2 by PCR; (5) the proportion of subjects with negative swabs for SARS-CoV-2 by PCR at Day 14; (6) the need for intubation and mechanical ventilation by Day 14; (7) the time to mechanical ventilation;

(8) the proportion of subjects, with at least one measurement of fever at baseline who were afebrile at Day 14; (9) mortality 30 days post-baseline.

The pre-defined safety endpoint of the study was to assess the safety and tolerability of opaganib. Details of the safety aspect evaluated in the study are provided below, under Safety evaluations.

The exploratory endpoint of the study was to assess the change from baseline in systemic markers of inflammation (D-dimer, cardiac troponin, C-reactive protein, lactate dehydrogenase and ferritin) by the end of treatment, as measured on Day 7 and Day 14.

As the majority of patients were discharged from the hospital without requiring supplemental oxygen before Day 14, and collection of oxygen ceased to be daily after Day 14, analyses based on total oxygen requirement (AUC) after Day 14 are not included. Additionally, all analyses based on measurements throughout the 14-day treatment period, including the secondary endpoints based on swabs for SARS-CoV-2 and those based on fever, the safety endpoint laboratory measurements, vital signs, and ECG, and the exploratory endpoint were not included for the following reason: the study was written at a time when patients were kept in the hospital until they had two consecutive negative viral swabs, and therefore, logistics had not been set up for collecting data that required physical measurement in the outpatient setting. Additionally, patients were encouraged to avoid coming to the hospital unless necessary for safety reasons. As most of the patients had been discharged before Day 14, these analyses could not be performed. More specifically, 68.2% of patients on opaganib and 50.0% of patients on placebo were discharged from hospital by Day 7, with proportions reaching 86.4% and 55.6% in the two groups, respectively, by Day 14. In addition to limiting our ability to analyze the Day 14 data due to limited sample numbers, these proportions also created negative bias that could confound the interpretation of a possible benefit with active treatment, as only those patients with worse outcomes remained hospitalized.

The post-hoc endpoints of the study included the following: (1) time to no longer receiving supplemental oxygen for at least 24 hours; (2) time to no longer receiving supplemental oxygen for at least 24 hours by Day 14 per standard of care (SoC) regimen of interest; (3) the percentage of subjects requiring intubation and mechanical ventilation by the end of safety follow-up; (4) time to intubation and mechanical ventilation; (5) mortality at the end of safety follow-up; (6) time to hospital discharge and percentage of patients discharged by Days 7, 14 and by the end of follow up; (7) time to at least two points improvement in the World Health Organization (WHO) Ordinal Scale (1) by Day 14. The WHO Ordinal Scale was not directly completed by the Investigators in the study. However, as all patients entered the study while hospitalized and requiring supplemental oxygen (score 4 or 5 on the scale, depending on oxygen supplementation), and the improvement sought in the study was that of discharge from hospital and no longer receiving supplemental oxygen (score 2), the improvement measured was that of at least two points on the scale. A detailed description of the WHO Ordinal Scale scores (points) and their definitions per patient condition is provided in Table S1.

**Table S1**: WHO Ordinal Scale used in the post-hoc analysis aimed at evaluating the time to two points improvement (1).

| WHO Ordinal Scale | Definition as per condition of patient |
|---|---|
| 0 | No clinical or virological evidence of COVID-19 infection |
| 1 | Ambulatory with no limitation of activities |

| | |
|---|---|
| 2 | Ambulatory with limitation of activities |
| 3 | Hospitalized, mild disease, no supplemental oxygen was required |
| 4 | Hospitalized, mild disease, low flow supplemental oxygen (nasal cannulas) |
| 5 | Hospitalized, severe disease, non-invasive positive pressure ventilation or |
| 6 | Intubation and mechanical ventilation used |
| 7 | Intubation and mechanical ventilation plus additional organ support measures |
| 8 | Death |

### 3. Statistical considerations

The results of the study were planned to be interpreted by the overall pattern of benefits of opaganib, based on numerical comparisons only. Descriptive statistics for continuous variables include n, mean, standard deviation, or median, minimum, and maximum, while descriptive statistics for categorical variables include patient counts and percentages.

Efficacy analyses

Efficacy analyses were performed in the mITT population including 22 patients in the opaganib and 18 patients in the placebo groups, with the following exceptions: for the post-hoc primary endpoint as well as the analysis of the time to 50% reduction from baseline in supplemental oxygen requirement, one patient randomized to the opaganib group was excluded from the mITT population due to no longer requiring supplemental oxygen prior to starting treatment, resulting in a total of 21 patients in the opaganib group. Additionally, a post-hoc sensitivity analysis was performed using the mITT population to investigate the potential impact of the imbalance of oxygen requirements at baseline on the secondary efficacy analyses. Two patients (both on placebo) who required greater than 40 L/min oxygen flow at baseline were removed from the post-hoc analysis, reducing the median oxygen flow in the placebo arm to 5.5 L/min while the opaganib arm remained unchanged at 6 L/min. Key secondary endpoints were analyzed using the resultant subject cohort. Kaplan-Meyer analysis was used to estimate cumulative incidence to events of interest, including time to 50% reduction from baseline in supplemental oxygen requirement based on oxygen flow, no longer requiring supplemental oxygen for at least 24 hours (including per SoC of interest), intubation and mechanical ventilation, death, hospital discharge, and time to at least two-point improvement on the WHO Ordinal Scale. In analysis of time to a favorable outcome death was taken as censored observation at the end of endpoint timeframe, as the worst possible result. Baseline was defined for each patient as the last available, valid, non-missing assessment before first study drug dose.

Safety evaluations

Safety analyses were performed in the safety population including 23 in opaganib and 18 patients in the placebo groups. Of note, one patient assigned to placebo, who mistakenly received opaganib on two days, was counted in both the placebo and opaganib populations in safety evaluations. The safety aspect evaluated in the study was the tolerability of opaganib, as determined by the following: (1) incidence rates of all treatment emergent AEs (TEAEs) and serious TEAEs throughout the duration of the study, including the follow-up period; (2) evaluation of vital signs; (3) evaluation of laboratory parameters (chemistry and hematology); (4) evaluation of ECG.

All adverse events (AEs) were coded using the Medical Dictionary for Regulatory Activities (MedDRA Version 23.0), using System Organ Class and Preferred Term classifications. Severity was graded according to the revised National Cancer Institute Common Terminology Criteria for AEs (NCI-CTCAE Version 5.0). For AEs that were not listed in the NCI-CTCAE Version 5.0, the physicians used the terms: mild/Grade 1, moderate/Grade 2, severe/Grade 3, life-threatening/Grade 4, or death/Grade 5 to describe the maximum intensity of the AE.

### 4. Randomization

A sample size of approximately 40 eligible patients in this study was judged adequate for the preliminary evaluation objectives rather than having been chosen for statistical consideration. Forty-two eligible patients were enrolled and were randomized using a 1:1 assignment ratio to treatment with either opaganib or placebo. There was a slight deviation from the 21/21 assignment, which resulted from the dynamic randomization applied without blocking. Three stratification factors were used in the minimization algorithm: (a) age at screening, ≥70 years of age, (yes or no), (b) HbA1c at screening, ≥6.5, (yes or no), and (c) oxygen requirement at baseline, requiring non-invasive positive pressure ventilation (e.g., via positive end-expiratory pressure, bilevel positive airway pressure, continuous positive airway pressure, high-flow nasal cannula), (yes or no). High-flow nasal cannulas were considered equivalent to positive pressure ventilation. The minimization algorithm used in the randomization was programmed by Bioforum Ltd. using the Viedoc system and applying the Pocock and Simon methodology (2), with biased coin probability of 0.8 and 'Range' as the variation method. Once the enrollment of a patient was approved, the Investigator assigned the patient an ID number created by the Viedoc system.

### 5. Treatment compliance

Compliance with the prescribed study treatment was assessed per subject, continuously from treatment initiation to end of treatment, with the drug accountability log of returned study bottles used to evaluate the number of capsules used, unused and lost. Additionally, the Dose

Missed/Reduced Log, which captured daily all missed or modified doses and reasons for modification, was used to assess compliance more accurately. Compliance rates are presented as percentages, with subject-level compliance rates computed as ([actual number of twice-daily doses taken] ÷ [expected number of twice-daily doses taken]) x 100.

**Supplementary data References**


1. WHO R&D Blueprint. COVID-19 Therapeutic Trial Synopsis. 2020;
2. Pocock SJ, Simon R. Sequential treatment assignment with balancing for prognostic factors in the controlled clinical trial. *Biometrics* 1975;31:103–115.
